# Acquired neutralizing breadth against SARS-CoV-2 variants including Omicron after three doses of mRNA COVID-19 vaccination and the vaccine efficacy

**DOI:** 10.1101/2022.01.25.22269735

**Authors:** Koichi Furukawa, Lidya Handayani Tjan, Yukiya Kurahashi, Silvia Sutandhio, Mitsuhiro Nishimura, Jun Arii, Yasuko Mori

## Abstract

To investigate the induction of neutralizing antibodies against Omicron after two and three vaccine doses in recipients of different ages. Physicians at Kobe University Hospital who had received the second dose of the BNT162b2 mRNA vaccine. At 2 months after the second vaccinations, the positive rate of neutralizing antibody against Omicron was 28%, and the titer was significantly lower than those against other variants, 11.8-fold and 3.6-fold lower than those against D614G and Delta, respectively. Unlike Delta, that positive rates of neutralizing antibody against Omicron were low in all age groups, and there was no significant difference in titers among age groups. Seven months after the 2nd dose, the positive rate of neutralizing antibody against Omicron decreased to 6%, but after the booster,3rd vaccination, it increased to 100%, and the titer was much higher than those at 2 and 7 months post-vaccination, 32-fold and 39-fold respectively. The booster vaccination effect was also observed in the younger at 41-fold, middle-aged at 43-fold, and older at 27-fold groups compared to the 7-month titers. Surprisingly, higher-than-predicted titers of the neutralizing antibodies against Omicron were induced after the booster vaccination regardless of recipient age, while this effect was not observed after two doses, indicating the induction of antibodies against common epitopes by the booster vaccination. Three doses can be confidently recommended to suppress the pandemic.

## Introduction

The coronavirus disease 2019 (COVID-19) pandemic declared by the World Health Organization (WHO) in March 2020 continues to affect all countries around the world. Based on the WHO COVID-19 dashboard as of 20 January 2022, there have been more than 332 million confirmed cases with more than 5.5 million deaths worldwide.^1^ Currently, the Omicron variant (B.1.1.529) of the original severe acute respiratory syndrome coronavirus 2 (SARS-CoV-2) is rapidly spreading and has become the dominant variant all over the world, including Japan.^2-6^ In efforts to control the pandemic, several vaccine platforms have been developed using the original SARS-CoV-2 virus (Wuhan-1) as the template, and the BNT162b2 mRNA vaccine (Comirnaty^®^, BioNTech-Pfizer, Mainz, Germany/New York, U.S.) was granted an emergency use authorization by the U.S. Food and Drug Administration (FDA) in December 2020, after showing high effectiveness in preventing infection.^7^

However, since April 2020, a variant bearing a D614G mutation of the spike protein and several SARS-CoV-2 variants classified as variants of concern (VOC) or variants of interest (VOI) by the WHO have emerged and spurred the spread of infection worldwide. As of this manuscript’s writing, SARS-CoV-2 variants classified as VOCs include Alpha (B.1.1.7) first detected in the UK,^8-10^ Beta (B.1.351) from South Africa,^11-14^ Gamma (P.1) from Brazil,^15,16^ Delta (B.1.617.2) from India,^17^ and Omicron (B.1.1.529) from South Africa and Bostwana, which is the latest VOC (detected in early November 2021).^18,19^ The emergence of these variants poses a tremendous challenge because the vaccine efficacy is negatively impacted by these variants.

In particular, the Beta and Gamma variants carry the E484K mutation in the receptor binding domain (RBD) of the spike protein, which allows evasion of neutralizing antibodies against the original SARS-CoV-2, further compromising the currently available therapy.^20-22^ The Delta variant has L452R, T478K, and P681R mutations in the RBD, which increases the virus transmissibility[—[leading to a higher viral load in infected individuals.^23,24^ However, the novel Omicron variant is the most mutated variant, with over 30 mutations in the spike protein and 15 mutations in the RBD.^25^ There is thus great concern about the Omicron variant’s higher transmissibility and immune escapes from COVID-19 vaccines and antibody-based therapies.^26-29^ It has also been demonstrated that vaccine-induced neutralizing antibody responses decrease with increased time post-vaccination.^30,31^

To counteract the decline of the neutralizing antibody response over time and the emergence of new variants including Omicron, a third dose of vaccination, which has been called a “booster vaccination,” has been approved and administered to individuals who were vaccinated >6 months ago; the booster vaccination has been shown to effectively induce a high titer of neutralizing antibody.^32^ However, it is unclear whether the effective neutralizing antibody against Omicron can be obtained by two or three doses of vaccination compared to the other variants. It is also not yet clear whether the characteristics of the vaccine’s recipients or side effects are related to the acquired immunity.

Here, we investigated the neutralizing breadth of sera against D614G, Alpha, Beta, Gamma, Delta, Kappa, and Omicron variants which were obtained from 82 vaccinated physicians without a history of SARS-CoV-2 infection, and we analyzed the correlations between the neutralizing antibody titers and the recipients’ background and side effects. We also followed and analyzed the neutralizing antibody titers against D614G, Delta, and Omicron in individuals after they received the booster (third dose of) vaccination.

## Participants and Methods

### Participant recruitment

From June 2021 to July 2021, blood samples from 82 physicians at Kobe University Hospital (Kobe, Japan) who had already received the second vaccination of the BNT162b2 mRNA COVID-19 vaccine and had never tested positive for SARS-CoV-2 infection were collected and analyzed (1st trial). As the 2nd trial, blood samples were collected again from the participants during the period from October to November 2021. In a 3rd trial, blood samples from the 72 participants who received booster vaccination were collected in January 2022 and analyzed. The sera of 24 unvaccinated and healthy adults were also collected and confirmed to have no antibody against SARS-CoV-2;^33^ these sera were used as the negative control group. This study was carried out after written consent was obtained from all participants. No statistical methods were used to predetermine the sample size.

### Questionnaire about side effects

When blood samples were collected, a questionnaire was used to obtain the participants’ background characteristics and information about any side effects that occurred within the first week after the first, second, and booster vaccinations. The contents of the questionnaire included age, sex, symptoms after vaccination such as fever (>37.5°), general fatigue, injection site pain (0: none, 1: mild, 2: moderate, 3: severe), the side of the injected arm, and others. The degree of injection site pain was subjectively determined by each participant.

### Measurement of neutralizing activity against SARS-CoV-2

The neutralization assay was performed using authentic viruses as recently described.^34,35^

### Preparation of SARS-CoV-2 variants

We used the SARS-CoV-2 Biken-2 (B2) strain with a D614G mutation as a conventional variant (accession no. LC644163); it was provided by the BIKEN Innovative Vaccine Research Alliance Laboratories (Osaka, Japan). The other variants, i.e., Alpha (GISAID ID: EPI_ISL_804007), Beta (GISAID ID: EPI_ISL_1123289), Gamma (GISAID ID: EPI_ISL_833366), Delta (GISAID ID: EPI_ISL_2158617), Kappa (GISAID ID: EPI_ISL_2158613), and Omicron (GISAID ID: EPI_ISL_7418017) had been isolated and were provided by the National Institute of Infectious Disease, Japan (Tokyo). Each variant was confirmed by the cDNA sequence of its spike gene.

### Statistical analysis

GraphPad Prism software (ver. 8.4.3) and STATA software (ver. 14.2) were used for the statistical analysis and preparation of figures. The Friedman test was used to compare the neutralizing antibody titer among the seven variants. The Mann-Whitney U-test or Kruskal-Wallis test was used to compare the neutralizing antibody titer or age among the different groups of participants. If a significant difference was found by the Friedman test or Kruskal-Wallis test, Dunn’s multiple comparison test was then performed. Results were considered significant when p-values were <0.05 (two-tailed).

### Ethical approval

This study was approved by the ethical committee of Kobe University Graduate School of Medicine (approval code: B200200).

## Results

### Participant characteristics and side effects after the first, second, and third vaccine doses

The demographic characteristics of the participants are summarized in Table 1. The median (interquartile [IQR]) numbers of days between the second dose of vaccination and the collection of serum samples in the 1st and 2nd trials were 53 [46–59] and 199 [192–203], respectively (hereinafter referred to as “2 months” and “7 months” after the vaccination). The median number of days between the booster vaccination and the collection of serum samples in the 3rd trial was 16 days. Because most of the physicians working in the hospital were men, most (71 of the 82) participants were males (86.6%).

**Table 1:** Characteristics of participants and side effects of each dose of vaccination.

| **Characteristics** | **First trial<sup>a</sup>**<br>**(n = 82)** | **Second trial<sup>b</sup>**<br>**(n = 82)** | **Third trial<sup>c</sup>**<br>**(n = 72)** |
| --- | --- | --- | --- |
| Sex, No. (%) |  |  |  |
| Male | 71 (87) | 71 (87) | 61 (85) |
| Female | 11 (13) | 11 (13) | 11 (15) |
| Age, years |  |  |  |
| Median (IQR) | 44 (33-58) | 44 (33-58) | 44 (33-53) |
| Age group, No. (%) |  |  |  |
| ≤ 38 years | 31 (38) | 31 (38) | 31 (43) |
| 39-58 years | 32 (39) | 32 (39) | 27 (38) |
| ≥ 59 years | 19 (23) | 19 (23) | 14 (19) |
| Days from 2nd dose of vaccination, days (IQR) | 53 (46-59) | 199 (192-203) | 274 (269-278) |
| Days from booster vaccination, days (IQR) | NA<sup>d</sup> | NA<sup>d</sup> | 16 (14-21) |
| **Side effects** | **1st dose of vaccination**<br>**(n = 82)** | **2nd dose of vaccination**<br>**(n = 82)** | **Booster vaccination**<br>**(n = 72)** |
| Fever, No. (%) | 0 (0) | 16 (20) | 24 (33) |
| Fatigue, No. (%) | 4 (5) | 14 (17) | 45 (63) |
| Injection site pain, No. (%) | 73 (89) | 71 (87) | 65 (90) |
| mild | 30 (37) | 27 (33) | 15 (21) |
| moderate | 39 (48) | 38 (46) | 35 (49) |
| severe | 4 (5) | 6 (7) | 15 (21) |

<sup>a</sup> Samples were collected from June to July 2021
<sup>b</sup> Samples were collected from October to November 2021
<sup>c</sup> Samples were collected in January 2022
<sup>d</sup> NA, not applicable

The side effects reported by the participants after the first, second, and booster vaccination are also listed in Table 1. Side effects at the time of the booster vaccination were as follows: fever in 24 participants (33%), general fatigue in 45 (63%), and injection site pain in 65 (89%) (15 mild, 35 moderate, and 15 severe). The frequency of systemic reactions including fever and fatigue increased in order, with the booster vaccination being the most frequent, but that of injection site pain did not change. No participant had serious adverse events.

### Neutralizing activity against Omicron compared with the other variants at 2 months post-vaccination

Although the neutralizing antibody titer varied, all 82 sera had neutralizing activity against D614G and Alpha. In contrast, most (but not all) of the sera had neutralizing activity against the Beta, Gamma, Delta, and Kappa variants; neutralizing antibody positive rates against the respective variants were 87.8%, 95.1%, 92.7%, and 90.2% of the participants. Surprisingly however, only 28% of the participants had the neutralizing activity against Omicron (Fig. 1A). The comparison of the antibody titers for each variant revealed that the antibody titers against Omicron were significantly lower than those against the six other variants (Fig. 1B). The average neutralizing antibody titers against Omicron were 11.8-fold (95%CI: 9.9–13.9) lower that that against D614G, 7.3-fold (95%CI: 6.3–8.5)) lower than that against Alpha, 3.2-fold (95%CI: 2.6–3.8)) lower than that against Beta, 4.9-fold (95%CI: 4.1–5.9)) lower than that against Gamma, 3.6-fold (95%CI: 3.1–4.2)) lower than that against Delta, and 2.6-fold (95%CI: 2.2–3.0) lower than that against Kappa.

**Fig. 1.**
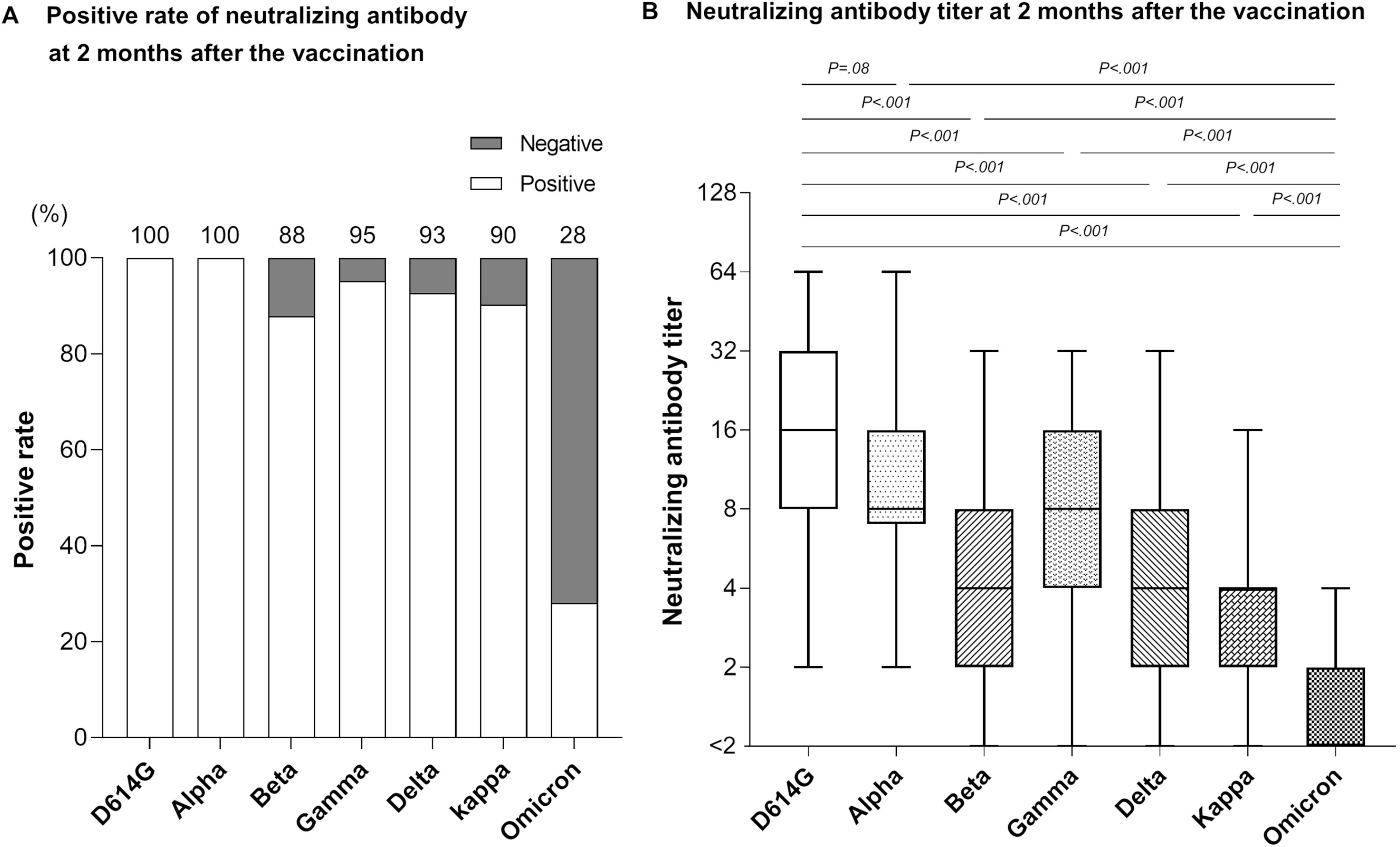
Neutralizing antibody against SARS-CoV-2 variants at 2 months post-vaccination. Sera of 82 recipients were tested for neutralizing activity against the SARS-CoV-2 variants D614G, Alpha, Beta, Gamma, Delta, Kappa, and Omicron. The neutralizing antibody titer is represented by the highest serum dilution that did not show any cytopathic effects. **(A)** The positive rate of neutralizing antibody against each variant. Sera with neutralizing activity at twofold dilution (the detection limit) or higher were considered positive. Numbers above the bar indicate the actual positive rate. (**B)** Box plot of the neutralizing antibody titers. The ends of the box represent the upper and lower quartiles, and the median is marked by the *horizontal line* inside the box. The ends of the ‘whiskers’ represent the minimum and maximum of all data. The titers against the seven variants were compared by the Friedman test and Dunn’s multiple comparisons test; two-tailed p-values were calculated.

The neutralizing antibody titer against D614G was significantly higher than those against Beta (3.6-fold; 95%CI: 3.0–4.3), Gamma (2.3-fold; 95%CI: 1.9–2.8), Delta (3.2-fold; 95%CI: 2.7–3.7), and Kappa (4.5-fold; 95%CI: 3.8–5.2).

### Neutralizing activity against Omicron and the other variants at 2 months after vaccination in each age group

We had divided the participants into three groups by age: the 31 participants (37.8%) who were ≤38 years old were classified as the younger group; the 32 participants (39.0%) aged 39–58 years old comprised the middle-aged group; and the 19 participants (23.2%) who were ≥59 years old constituted the older group (Table 1). Although all of the participants’ sera showed neutralizing activity against D614G and Alpha regardless of age, sera with negative neutralizing activity against other variants were observed in all age groups, and the negative proportion tended to be higher in the older group (Fig. 2A). The positive rate of sera with neutralizing antibody against Omicron was low in all three age groups; only 32% had the neutralizing antibody against Omicron even in the younger age (≤38 yrs) group. The comparison of neutralizing antibody titers among the three age groups showed results similar to the positive rate: the median [range] of neutralizing antibody titer against Delta was significantly lower in the older age group (3 [<2–8]) compared to the other two groups: (4[<2–32] vs. the younger group, p=0.03) and (4 [<2–32] vs. the middle-aged group, p=0.04). Similar results were also obtained for the median [range] of neutralizing titer against Kappa: (2 [<2–4] in the older group) versus (4 [<2–16] in the younger group, p=0.02) and (4 [<2–16] in the middle-aged group, p=0.05) (Fig. 2B).

**Fig. 2.**
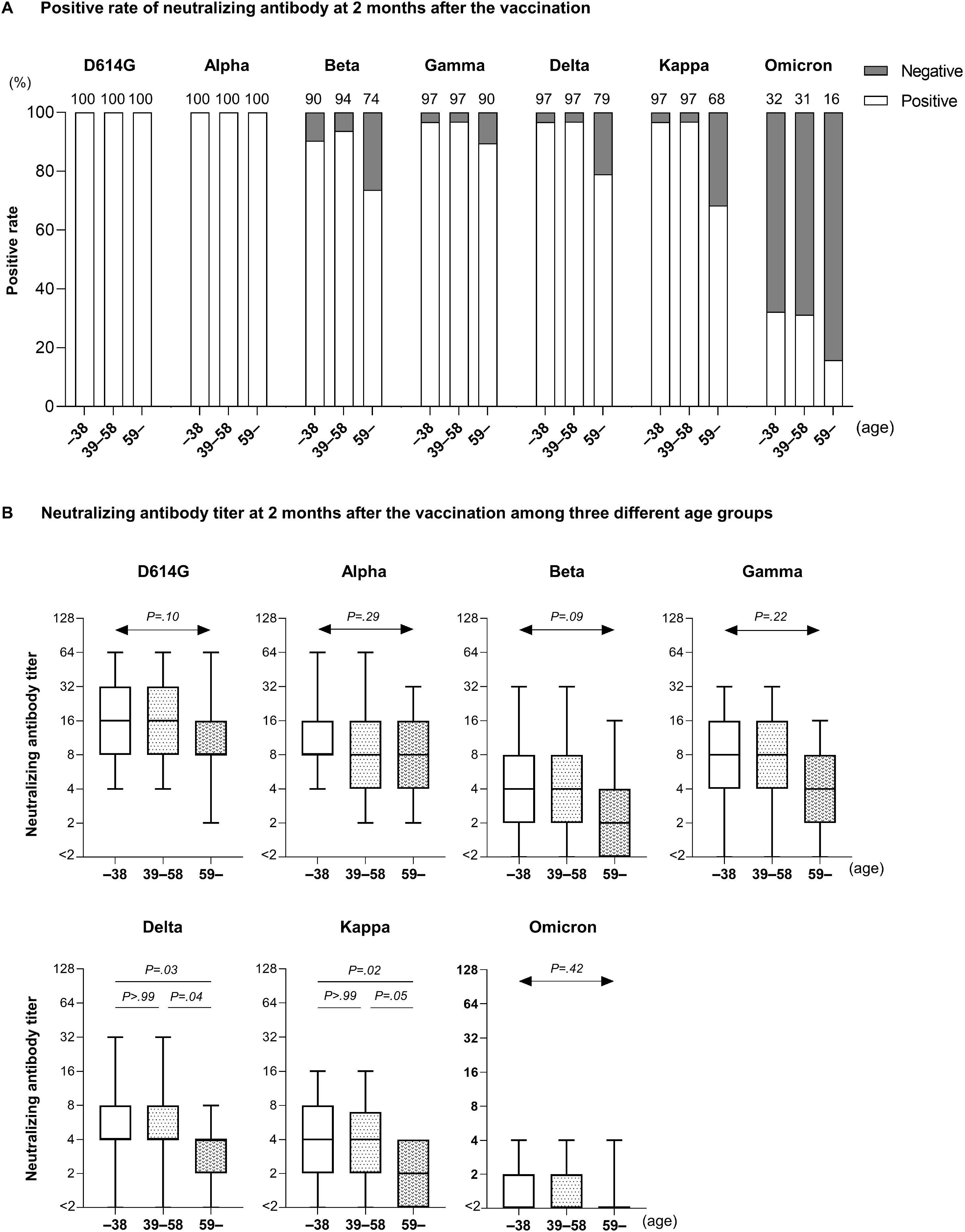
Comparison of neutralizing antibodies against SARS-CoV-2 variants among three age groups. The 82 participants were divided into three age groups: 31 participants (37.8%) who were ≤38 years old, 32 participants who were 39–58 years old, and 19 participants who were ≥59 years old, and the recipients’ sera were tested for neutralizing activities against the seven SARS-CoV-2 variants. (**A)** The positive rates of neutralizing antibodies against each variant in the three age groups shown. Sera with neutralizing activity at twofold dilution (detection limit) or higher were considered positive. Numbers above the bar are the actual positive rate. (**B)** Comparison of neutralizing antibody titers against each variant among the age groups. Ends of the box indicate the upper and lower quartiles, and the median is marked by the *horizontal line* inside the box. Ends of the ‘whiskers’ = the min. and max. of all data. The Kruskal-Wallis test and Dunn’s multiple comparison test were performed, and two-tailed p-values were calculated.

### Side effect-related differences in neutralizing activity and recipient characteristics

Our comparison of neutralizing antibody titers against any of the SARS-CoV-2 variants showed no significant difference in the percentages of participants with and without side effects, e.g., fever, injection site pain, and fatigue (eFig. 1A in Supplement). However, the comparison of participant ages and side effects revealed that the median age of the participants with fever was significantly younger than that of the participants without fever: 37 years (range 28–64 yrs) versus 46 (29–68) years, p=0.02. There were no significant between-group differences in the median age of the participants with or without fatigue (44 years [range 28–68 yrs] vs. 39 [28–64], p=0.97) or regarding the degrees of injection site pain (none: 41 [31–68] vs. mild: 50 [30–64] vs. moderate: 42 [28–64] vs. severe: 39 [36–51]) (eFig. 1B in Supplement).

### Neutralizing activity against three variants at 7 months after two vaccine doses and after the booster

We determined the positive rates of neutralizing antibody against D614G, Delta, and Omicron at 2 and 7 months after the two vaccine doses. At 7 months after the vaccination, 93% of the 82 participants had neutralizing antibody against D614G, 67% against Delta, and only 6% against Omicron (Fig. 3A). However, surprisingly, after the booster vaccination, all 72 participants who had received the booster had the neutralizing antibody against these three variants. Figure 3B illustrates the changes in the neutralizing antibody titers against these variants during the same period. The neutralizing antibody titers against D614G and Delta were significantly decreased at 7 months compared to those at 2 months after the second vaccination: D614G, 2.2-fold (95%CI: 1.9–2.5) lower; Delta, 2.1-fold (95%CI: 1.8–2.4) lower. Although the neutralizing antibody titers against Omicron also tended to decrease during this period, no significant difference was observed: median [range] neutralizing antibody titer was <2 [<2–2] at 2 months versus <2 [<2–4] at 7 months (p=0.54).

**Fig. 3.**
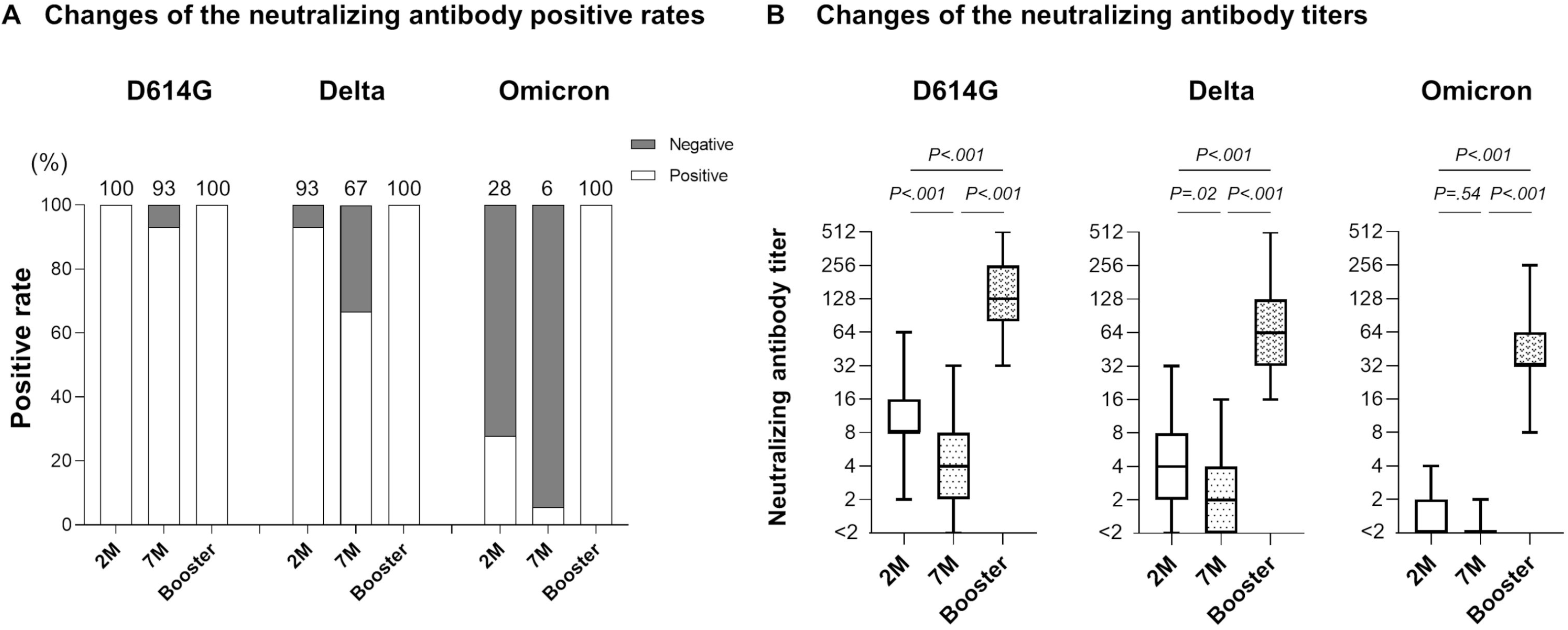
Neutralizing antibody after booster vaccination. Sera of the 72 participants who received the booster vaccination were tested for neutralizing activity against D614G, Delta, and Omicron at 2 and 7 months after the two doses of vaccination and after the booster (third) vaccination. **(A)** Changes in the neutralizing antibody positive rate against each variant. Sera with neutralizing activity at twofold dilution (detection limit) or higher were considered positive. Numbers above the bar: the actual positive rate. **(B)** Changes in the neutralizing antibody titers against each variant. Box ends: the upper and lower quartiles. The *horizontal line* inside the box: median. Whisker ends: the min. and max. of all data. The titers were compared by the Friedman test and Dunn’s multiple comparisons test; two-tailed p-values were calculated.

Notable, the neutralizing antibody titers after the booster vaccination were much higher than those at 2 and 7 months post-vaccination against all variants, as follows. D614G: 13-fold (95%CI: 11–16) and 31-fold (95%CI: 25–38) higher than that at 2 and 7 months, respectively; Delta: 16-fold (13–20) and 35-fold (95%CI: 29–43); Omicron: 32-fold (27–37) and 39-fold (95%CI: 32–46).

### Comparison of neutralizing antibody titers after booster vaccination in the three age groups

We compared the effects of booster vaccination on the neutralizing antibody titers in each age group (Fig. 4A–C). Seven months after the 2nd vaccination, the neutralizing antibody titers against D614G and Delta tended to be low in the older age group, and those against Omicron were much lower in all three age groups. However, the booster vaccination effectively increased the neutralizing antibody titers for all three variants including Omicron, regardless of the participants’ ages, as follows. The younger, middle-aged, and older groups values after the booster compared to the values at 7 months were D614G: 28-fold (22–36), 28-fold (95%CI: 18–42), and 47-fold (95%CI: 29–75) greater; those for Delta were 32-fold (95%CI: 23–44), 38-fold (95%CI: 26–55), and 39-fold (95%CI: 29–52) greater; and those for Omicron were 41-fold (95%CI: 30–56), 43-fold (95%CI: 32–58), and 27-fold (95%CI: 20–36) greater, respectively.

**Fig. 4.**
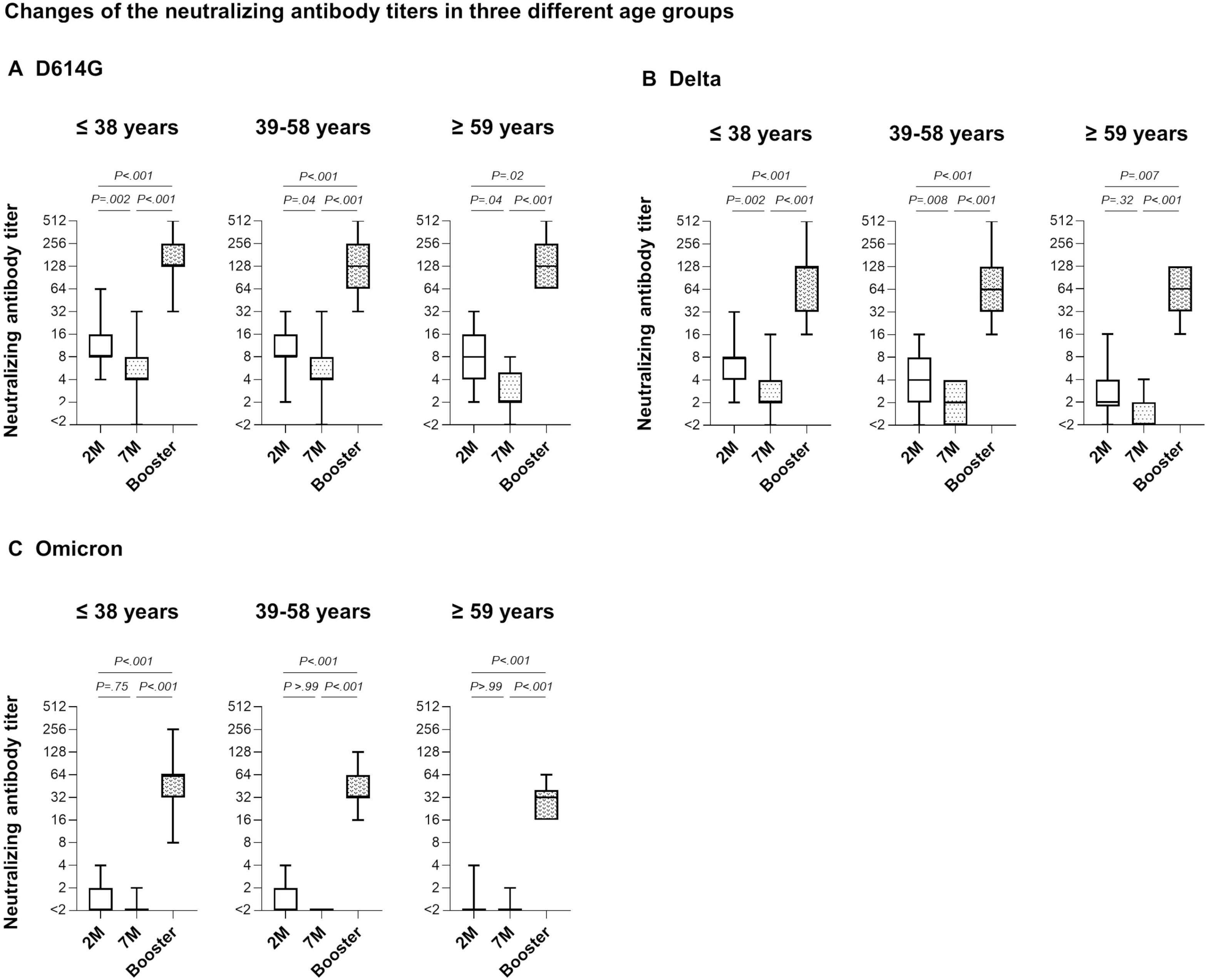
Changes in the neutralizing antibody titer after booster vaccination in the age groups. The 72 recipients who received the booster vaccination were divided into three age groups: 31 participants who were ≤38 years, 27 participants who were 39–58 years, and 14 participants who were ≥59 years. The neutralizing antibody titers against D614G **(A)**, Delta **(B)**, and Omicron **(C)** at 2 and 7 months after two doses of vaccination and after the booster vaccination were compared in each age group. Box ends: the upper and lower quartiles. The *horizontal line* inside the box: median. Whisker ends: the min. and max. of all data. The Friedman test and Dunn’s multiple comparisons test were performed; two-tailed p-values were calculated.

## Discussion

We evaluated neutralizing activities against authentic SARS-CoV-2 variants in the sera of adults vaccinated with two or three doses of the BNT162b2 mRNA vaccine. We also analyzed the changes in the neutralizing antibody titers after the booster vaccination in different age groups, and we evaluated the relationships between side effects, the participants’ ages, and the acquired neutralizing antibody titer. This is the first report on the detailed efficacy of two doses and the booster vaccination for a Japanese population using authentic viruses including Delta and Omicron.

Our results demonstrated that only 28% of the vaccine recipients had neutralizing antibodies against Omicron at 2 months after their second vaccination, and the neutralizing antibody titer against Omicron was significantly lower than those against all six of the other variants tested. This low positive rate of neutralizing antibody against Omicron was distinctly different from what we observed for other variants, in which 88%–100% of recipients had the neutralizing antibody against other variants at 2 months after the vaccination, suggesting that the neutralizing antibody against Omicron cannot be sufficiently induced by only two doses of vaccination, as has been noted in other reports.^28,29,36,37^

At 7 months after the second vaccination, that tendency was even more pronounced, with only 6% of the participants showing neutralizing antibodies against Omicron, suggesting a high risk of breakthrough infection or reinfection. Regarding the potential cause of this finding, it has been reported that mutations on the spike protein of Omicron, especially K417N, N440K, G446S, S477N, T478K, E484A, Q493R, G496S, Q498R, N501Y, and Y505H, were responsible for the variant’s broad evasion from antibodies.^38^ In case of the Delta variant, L452R, T478K, and P681R have been reported to promote the interaction between the spike and angiotensin-converting enzyme 2 (ACE2) receptor by inducing structural changes in the binding domain, resulting in immune evasion and increased transmissibility.^39-41^ In our present investigation, 66.7% of the participants had neutralizing antibodies against Delta at 7 months after their second vaccination, suggesting that neutralizing antibodies against Delta could be more easily induced by two doses of vaccination compared to Omicron.

We also observed that the booster vaccination induced much higher levels of neutralizing antibodies against Omicron compared to those after two vaccine doses. This may indicate that the booster vaccination can induce sufficient immunity against Omicron and the other variants that may appear in the future, by inducing neutralizing antibodies that recognize common epitopes, and by promoting affinity maturation of the neutralizing antibody.^42-44^

Our comparison of neutralizing antibody titers among the three age groups at 2 months after the second vaccination showed that neutralizing activity against all seven tested variants tended to be lower in the older participants (and the activity was significantly lower only for Delta and Kappa). However, our findings of no significant difference in the neutralizing antibody titers against Omicron among the present age groups and that only 32% of the participants in the youngest group had neutralizing antibodies reflects the universally low neutralizing activity for Omicron, unlike that for Delta.

However, the increase in the neutralizing antibody level induced by the booster vaccination did not show a large difference among the three age groups, and the neutralizing antibody against Omicron was sufficiently induced in all age groups, suggesting that the effect of the booster vaccination is guaranteed even in older individuals and that the booster should be recommended for all individuals regardless of age.

The proportion of participants who had fever or general fatigue after vaccination in this study increased from the first dose to the third dose of vaccination, whereas the distribution of injection site pain was almost the same across the doses, indicating that the vaccinations’ systemic side effects such as fever and general fatigue may increase mainly as a consequence of a stronger immunological response for the second and third doses.^45-47^ However, we did not detect a correlation between the neutralizing antibody titer induced by two doses of vaccination and vaccination-related side effects, indicating that fever and other side effects are not specific immune reactions to the viral spike protein, but rather may be specific to other materials contained in the vaccine. This relationship remains controversial, and further studies including a booster vaccination are necessary to evaluate it.^48^

Overall, our findings showed that a sufficient neutralizing antibody titer against Omicron could not be induced by only two vaccine doses, suggesting a high risk of breakthrough infection or reinfection. Moreover, the elderly may not be equally protected from Delta and other variants. Although systemic side effects such as fever and general fatigue may increase, our study revealed that the third dose of the vaccination effectively induced neutralizing antibodies against Omicron regardless of the recipients’ age. The booster should therefore be recommended to help prevent the further spread of the COVID-19 pandemic.

## Data Availability

All data produced in the present study are not available.

## Acknowledgments

We thank Kazuro Sugimura MD, PhD (Superintendent, Hyogo Prefectural Hospital Agency and Professor, Kobe University) for his full support to promote this study. We thank BIKEN Innovative Vaccine Research Alliance Laboratories for providing SARS-CoV-2 B2 strain. We thank the National Institute of Infectious Disease Japan for providing SARS-CoV-2 Alpha, Beta, Gamma, Delta, Kappa and Omicron variants.

## Authors’ Statement

All authors attest that they meet the ICMJE criteria for authorship.

## Presentation at meetings

This study has never been presented anywhere.

## Contributions

All authors contributed to the concept of this article. KF drafted the manuscript; YM provided revisions; KF, YK, JA, MN, and YM analyzed the data; KF, YK, LT, and SS did the experiments; YM supervised the experiments; KF collected the samples; YM conducted the project; All authors approved the final version of the manuscript.

## Figure legends

**eFigure 1.**
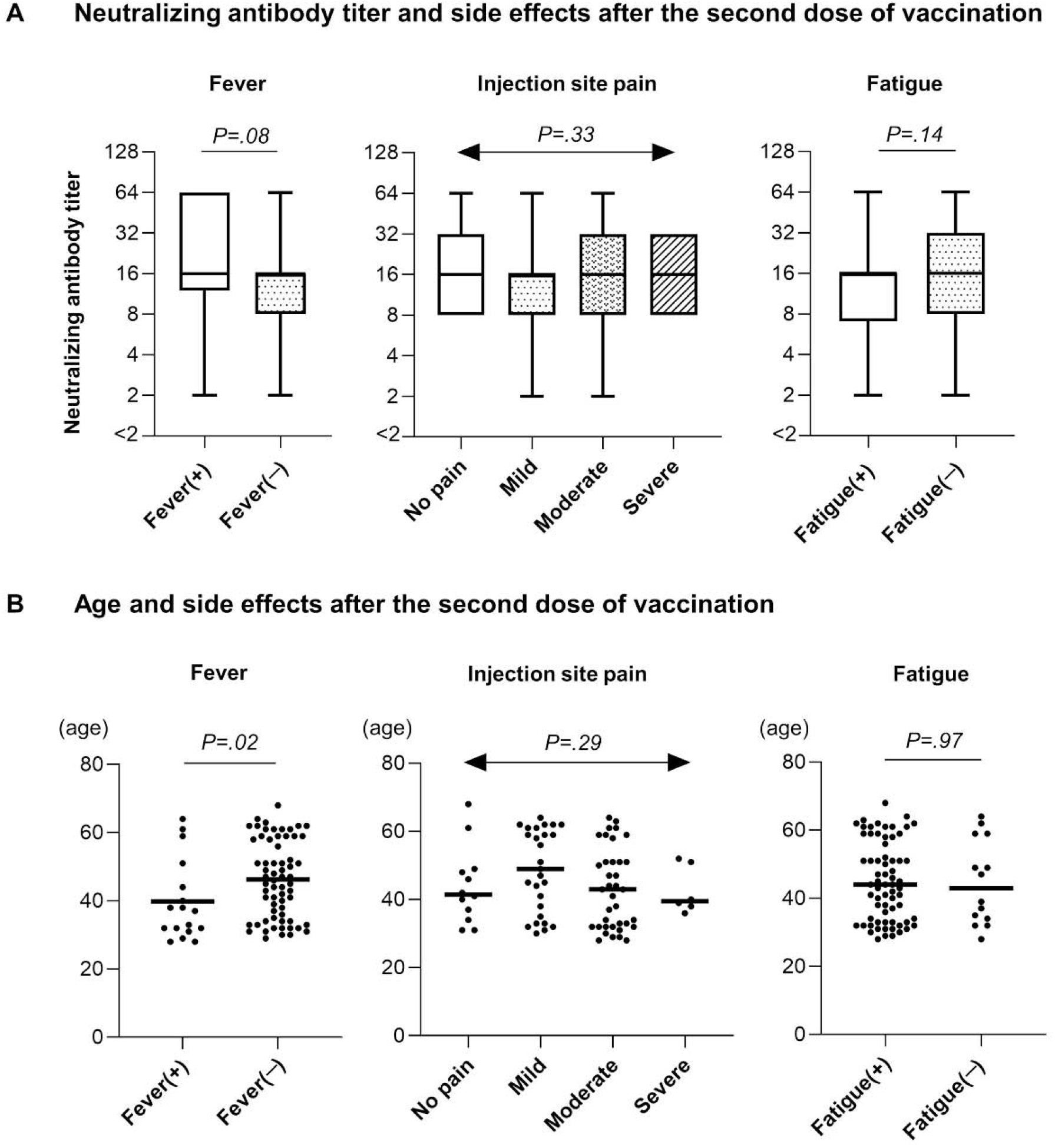
Comparison of side effects and neutralizing antibody titers or recipient’s age. Eighty-two recipients were divided according to the appearance of fever, fatigue, and the degree of injection site pain after the second dose of vaccination. The neutralizing antibody titers **(A)** and age of recipients **(B)** were compared by each group. The ends of e box show the upper and lower quartiles, and the median is marked by the horizontal line inside the box. The ends of the “whiskers” represent the minimum and maximum of all data. Lines in eFigure 1B indicate the mean age of recipients in each group. Mann-Whitney U test was performed for the comparison by the presence or absence of fever and fatigue, and Kruskal-Wallis test was performed for the comparison by the degree of injection site pain. Two-tailed p-values were calculated.

